# Neural Indicators of Motor and Cognitive Functioning in Sarcopenia Using Functional Near-Infrared Spectroscopy

**DOI:** 10.64898/2026.03.04.25342448

**Authors:** Bora Mert Şahin, Murat Kara, Kübra Erdoğan, Mahmut Esad Durmuş, Özgür Kara, Bayram Kaymak, Aykut Eken

## Abstract

Sarcopenia is a geriatric condition characterized by the loss of muscle strength, muscle mass, and physical performance, yet its neural mechanisms remain insufficiently understood. This study aimed to identify cortical indicators of motor and cognitive functioning in individuals with sarcopenia using functional near-infrared spectroscopy (fNIRS), along with electromyography (EMG) and hand dynamometer measurements. 30 sarcopenia patients (age 67.33 ± 7.48, F/M: 22/8) and 38 healthy controls (age 65.37 ± 4.18, F/M: 29/9) participated in three experimental sessions designed to probe different neural systems: a Hand Grip task to assess motor function, an N-Back task to evaluate working memory, and an Oddball task to measure attention and inhibitory control. fNIRS measurements were carried out during all experimental sessions, while EMG and force output were extracted from the Hand Grip task. Group differences and neural-behavioral relationships were examined using t-tests, correlations, and repeated measures analyses.

Participants with sarcopenia demonstrated significantly reduced EMG activity and force production. Although motor cortex responses during the Hand Grip task were similar between groups, the N-Back task revealed lower activation in the precentral, middle frontal, and superior frontal regions in the sarcopenia group. In contrast, the Oddball task showed increased right-hemisphere activation in sarcopenic individuals, suggesting compensatory recruitment. Significant correlations between cortical activity, grip strength, and Chair Stand Test performance indicated shared neural pathways linking motor and cognitive function.

These findings highlight altered neural processing in sarcopenia and emphasize the importance of integrating neuroimaging with clinical measures to advance early detection and targeted intervention strategies.

**Highlights:**

- fNIRS assessed motor and cognitive cortical activity in sarcopenia.
- Sarcopenia showed lower EMG amplitude and grip force output.
- No group difference in motor cortex activation during hand grip.
- N-back revealed lower frontal and precentral activation in sarcopenia.
- Oddball showed higher right-hemisphere activation in sarcopenia.

## 1. Introduction

Aging can lead to various challenges that negatively impact the quality of life of people and can be characterized by a gradual decline in biological and physiological functions (Gilbert, 2000). It is a complex process that involves multiple mechanisms that begin at the cellular level and affects the entire organism over time. These mechanisms mainly result in a reduction in bodily functions and cause difficulties during daily activities. Such challenges can cause mobility limitations in the elderly populations, which can increase the risk of injury, hospitalization, and mortality. Given the growing elderly population worldwide, the identification, prevention, and treatment of diseases associated with aging are becoming critical for public health (Grinin et al., 2023). Among the conditions that adversely affect the life quality in older adults, the decline in muscle mass and strength is particularly significant which is also highly associated to cognitive decline due to sharing pathophysiological pathways (Sui et al., 2020). In this context, sarcopenia, a condition directly linked to the loss of muscle mass and strength, has emerged as a significant health concern.

Sarcopenia is a term coined by Rosenberg in 1989 and it derives from the Greek words “sarx” (flesh) and “penia” (loss) (Baumgartner et al., 1998). Sarcopenia is a widespread condition among individuals over the age of 60, with a prevalence rate of 11% in men and 9% in women globally (Papadopoulou et al., 2020) and characterized by a significant loss of skeletal muscle mass, strength, and function in older individuals (Cruz-Jentoft et al., 2010). While initially defined solely as the loss of muscle mass, recent studies have highlighted that the declines in muscle strength and function are also key indicators for diagnosing sarcopenia (Clark and Manini, 2008). Muscle function in the elderly can be assessed through voluntary tests such as maximal isometric or dynamic contractions, and involuntary methods like electrically stimulated twitches or tetanic responses, which help isolate muscular from neurological factors affecting performance (Hunter et al., 1998). In individuals over the age of 45, reductions in muscle function can significantly lower quality of life, crossing a critical threshold that increases the risk of disability and mortality. In such cases, the diagnosis of sarcopenia is often made (Janssen et al., 2002). Recent studies have also highlighted the importance of assessing regional muscle groups, particularly the anterior thigh, and integrating functional and neuromotor evaluation for earlier and more accurate detection of sarcopenia, especially in rehabilitation contexts (Kara et al., 2021).

Sarcopenia was initially thought as the natural decline in muscle mass that occurs with aging (Rosenberg, 1997). However, recent advancements in sarcopenia research have revealed that this condition extends beyond mere muscle mass loss, encompassing significant reductions in muscle strength and function as well (Clark and Manini, 2008). Recognizing this, the European Working Group on Sarcopenia in Older People (EWGSOP) introduced revised criteria that underscore the importance of evaluating both muscle strength and physical performance when diagnosing sarcopenia (Cruz-Jentoft et al., 2010). Despite these updates, the exact mechanisms driving sarcopenia remain debated, with no clear consensus on the most influential factors in its onset and progression (Jones et al., 2009). Emerging research points to the potential role of neurological factors, particularly the degeneration of motor neurons and the disruption of neuromuscular junctions, as key contributors to the weakening of muscles and the overall decline in physical function seen in older adults (Moreira-Pais et al., 2022). These findings suggest that while sarcopenia is a muscular issue, it also has critical neurological dimensions that require further exploration.

The relationship between sarcopenia and cognitive functions has emerged as a key area of research. Studies and meta-analyses have supported the notion that sarcopenia can impact cognitive abilities, although the neurological mechanisms behind these effects remain poorly understood (Beeri et al., 2021). In a community-dwelling sample, sarcopenic individuals had over six times the odds of combined physical and cognitive impairment, as measured by MoCA (Montreal Cognitive Assessment) and AD8 (Ascertaining Dementia 8), compared with non-sarcopenic controls (Tolea and Galvin, 2015). Moreover, poor performance on the five-times CST (Chair Stand Test), an established functional marker of lower-limb strength, was associated with roughly 2.2 times higher odds of mild cognitive impairment and with deficits in attention, executive function and processing speed in Japanese elders (Makizako et al., 2022). Notably, patients with sarcopenia have been found to experience a loss of grey matter in cortical regions, which is associated with declines in both motor and cognitive functions. However, the underlying neurological pathways that link these impairments have yet to be fully elucidated (Yu et al., 2020). As such, future research should aim to explore the neurological changes in sarcopenia patients in greater detail, particularly focusing on how these changes contribute to both motor and cognitive deterioration. Understanding this connection could provide deeper insights into the broader impacts of sarcopenia on brain health and function. To understand structural and functional changes induced by sarcopenia, neuroimaging techniques were previously used.

Researchers have employed both electrophysiological and neuroimaging techniques to investigate the neurological underpinnings of the condition. For instance, studies combining EEG and EMG have demonstrated heightened corticomuscular coherence in sarcopenic individuals, suggesting a compensatory increase in synchronization between the motor cortex and spinal motor neurons to counteract muscle weakness (Gennaro et al., 2020). Moreover, MRI investigations have revealed structural brain alterations in sarcopenia, such as reduced cortical thickness in the superior parietal and precuneus regions, diminished left hippocampal volume, and lower periventricular white matter density (Kim et al., 2022). Recent large-scale studies, such as Gurholt et al. (2024), have further supported these findings by demonstrating widespread reductions in cortical thickness, brain volumes and white matter integrity among individuals with probable sarcopenia, highlighting structural brain changes as significant mediators between sarcopenia and cognitive decline (Gurholt et al., 2024). Additionally, Kim et al. (2024) developed a multimodal neuroimaging-based prediction model, revealing distinct relationships between muscle mass, strength and function with cortical atrophy, amyloid-beta retention and white matter hyperintensities in older adults with sarcopenia (Kim et al., 2024). Complementing these findings, Trost et al. (2023) performed an fMRI research showing that during dual-task paradigms involving both motor and cognitive components, sarcopenic subjects exhibit increased activation in premotor and prefrontal cortices compared to healthy controls, indicating the recruitment of additional neural resources under challenging conditions (Trost et al., 2023). Furthermore, Teo et al. (2024) demonstrated that deep-learning-derived neuroimaging biomarkers, specifically reduced temporalis and sternocleidomastoid muscle volumes, independently predicted poor functional outcomes in stroke patients undergoing endovascular thrombectomy, underscoring the interplay between muscular and neurological health in sarcopenic conditions (Teo et al., 2024). Among these techniques, functional Near-Infrared Spectroscopy (fNIRS) is an emerging neuroimaging technique that enables non-invasive, portable and cost-effective measurement of cortical activity by detecting changes in oxygenated and deoxygenated hemoglobin (HbO, HbR) levels in the brain. Its capacity to assess brain function in naturalistic environments makes it particularly suitable for clinical use, especially in elderly or mobility-impaired populations (Irani et al., 2007). fNIRS has been applied across a range of neurological and psychiatric conditions such as Alzheimer’s disease, Parkinson’s disease. fNIRS can offer insights into disease-specific cortical dysfunction (Arenth et al., 2007).

In rehabilitation and geriatric settings, fNIRS can be especially valuable due to its tolerance to motion artifacts and easy integration into clinical tasks, which makes fNIRS a great tool for monitoring neural responses during motor or cognitive exercises used in physical therapy (Bunce et al., 2006). Recent fNIRS applications include investigating populations with cognitive impairment, which shows its usefulness for aging-related research (Yang et al., 2019).

In this study, we examined cortical regions encompassing the Precentral Gyrus (PcG), which contains the primary motor cortex involved in voluntary movement planning and execution, as well as three frontal regions: the Middle Frontal Gyrus (MFG) and Superior Frontal Gyrus (SFG), serving as key nodes in working memory, attention and executive control, and Inferior Frontal Gyrus (IFG), associated with response inhibition and high-order executive control (Banker and Tadi, 2025; El-Baba and Schury, 2025; Boisgueheneuc et al., 2006). Three experimental tasks were administered: Hand Grip, N-Back and Oddball paradigm. Consistent with prior neuroimaging findings, the Hand Grip task was expected to elicit activation in the PcG region, the N-Back task to engage MFG/SFG as components of dorsolateral prefrontal cortex implicated in working memory, and the Oddball paradigm to recruit IFG region due to its involvement in inhibitory control and higher-order cognitive processing (Noble et al., 2011; Suzuki et al., 2018; Bae et al., 2024). To understand the cortico-muscular relationship in sarcopenia patients, we also utilized Electromyography (EMG) during Hand Grip task. Unveiling the relationship between motor and cognitive functions in sarcopenia might open new avenues for both clinical and fundamental research. These findings could enhance diagnosis, treatment strategies and deepen our understanding of the broader neurological underpinnings of aging-related muscle loss.

## 2. Methods

### 2.1. Participants

This study was approved by the Human Research Ethics Committee of TOBB University of Economics and Technology (Approval number 2022-25). The study included patients diagnosed with sarcopenia and healthy individuals without the condition, selected from among people over the age of 60 who visited the Geriatrics Clinic at Dr. Abdurrahman Yurtaslan Ankara Oncology Hospital. Before the experiment, geriatric doctors (K.E., Ö.K., M.E.D.) collected data on participants’ age, height, weight, vitamin D levels, hand grip strength (HGS), Chair Stand Test (CST), and Mini Mental State Examination (MMSE). Participants with grip strength below the threshold (HGS < 19 kg for women, HGS < 32 kg for men) were diagnosed with sarcopenia.

A total of 95 participants, including 75 women and 20 men, took part in the experiments. After excluding participants who could not complete the experiment, did not meet the required literacy criteria, or whose signal quality was insufficient, 68 participants remained in the study. This included 51 women (29 healthy controls, 22 with sarcopenia) and 17 men (9 healthy controls, 8 with sarcopenia), resulting in a total of 38 healthy controls and 30 sarcopenic individuals. The details of the participants included in the study are provided in Table 1.

### 2.2. Experimental Design

In this study, three different experiments were conducted to measure participants’ motor and cognitive activities: hand grip, N-Back, and Oddball experiments. The experimental design was implemented using the PsychoPy application, and synchronization between the stimuli from the experiment computer and the fNIRS recordings on the measurement computer was ensured via Lab Streaming Layer (LSL) (Peirce et al., 2019). Before starting the experiments, each participant signed an informed consent form in accordance with Helsinki declaration (World Medical Association, 2013).

**Table 1:** Subject Information and t-test results.

| Variable | Control Mean<br>± SD | Sarcopenic<br>Mean ± SD | t-test |
| --- | --- | --- | --- |
| Age | $65.37 \pm 4.18$ | $67.33 \pm 7.48$ | $t(66) = -1.371$,<br>$p = 0.175$ |
| Menopause<br>Age | $47.68 \pm 6.19$ | $48.14 \pm 6.53$ | $t(66) = -0.253$,<br>$p = 0.801$ |
| Height | $160.55 \pm 9.53$ | $157.93 \pm 9.97$ | $t(66) = 1.103$,<br>$p = 0.274$ |
| Weight | $81.45 \pm 15.55$ | $77.13 \pm 14.11$ | $t(66) = 1.183$,<br>$p = 0.241$ |
| Quadriceps | $34.78 \pm 8.37$ | $31.40 \pm 6.96$ | $t(66) = 1.778$,<br>$p = 0.114$ |
| Grip<br>Strength | $27.17 \pm 8.69$ | $19.67 \pm 5.77$ | $t(66) = 4.065$,<br>$p < 0.001^{***}$ |
| CST | $10.39 \pm 2.37$ | $11.73 \pm 3.82$ | $t(66) = -1.763$,<br>$p = 0.083$ |
| Vitamin D | $18.83 \pm 12.61$ | $21.65 \pm 15.46$ | $t(66) = -0.757$,<br>$p = 0.452$ |
| MMSE | $28.79 \pm 1.23$ | $28.52 \pm 1.68$ | $t(66) = 0.765$,<br>$p = 0.447$ |

Notes: \* $p < 0.05$ , \*\* $p < 0.01$ , \*\*\* $p < 0.001$ .

In a quiet room, participants were seated in front of the experiment computer. Before each experiment, the procedure was explained to the participants. Then, to help them understand the experiment flow, a trial run without data collection was conducted before the experiment. Once it was confirmed that the participant understood the procedure and felt comfortable, data collection began. In the beginning of each experiment, participants were asked to remain still for 10 seconds to record baseline values.

In the first experiment, hand grip, participants were instructed to squeeze and release the hand dynamometer in their right hand according to the instructions displayed on the screen. After a 20-second rest period, participants were prompted with a “Prepare the dynamometer” message for 5 seconds, followed by “Squeeze” and “Release” cues on the screen. They were asked to squeeze for 2 seconds and release for 1 second, repeating this cycle for 35 seconds, a total of 10 times.

The second experiment is the N-Back task, commonly used to assess memory function (Owen et al., 2005; Keles et al., 2017). The N-Back task includes two conditions: 0-Back and 2-Back. In the 0-Back condition, participants press the space bar on the experiment computer when they see the letter “X” among a sequence of random letters. In the 2-Back condition, participants must remember the sequence of letters and press the space bar if the current letter matches the one shown two letters prior. For both tasks, a 10-second instruction screen is shown, followed by a sequence where each letter appears for 2.5 seconds, interspersed with a blank screen for 0.5 seconds.

The final experiment is the Oddball task, designed to assess attention function (Kim, 2014). The Oddball task includes two conditions: standard stimulus and Oddball stimulus. In the standard stimulus condition, the letter “O” appears on the screen for 1.5 seconds, followed by a blank screen for 0.2 seconds. Participants are instructed to press the “O” key whenever they see the letter “O” on the screen. In the Oddball stimulus condition, the letters “X” and “O” appear in a random sequence, and participants press the corresponding key based on the letter shown. Block diagrams for each experiment are given in Figure 1.

**Figure 1:**
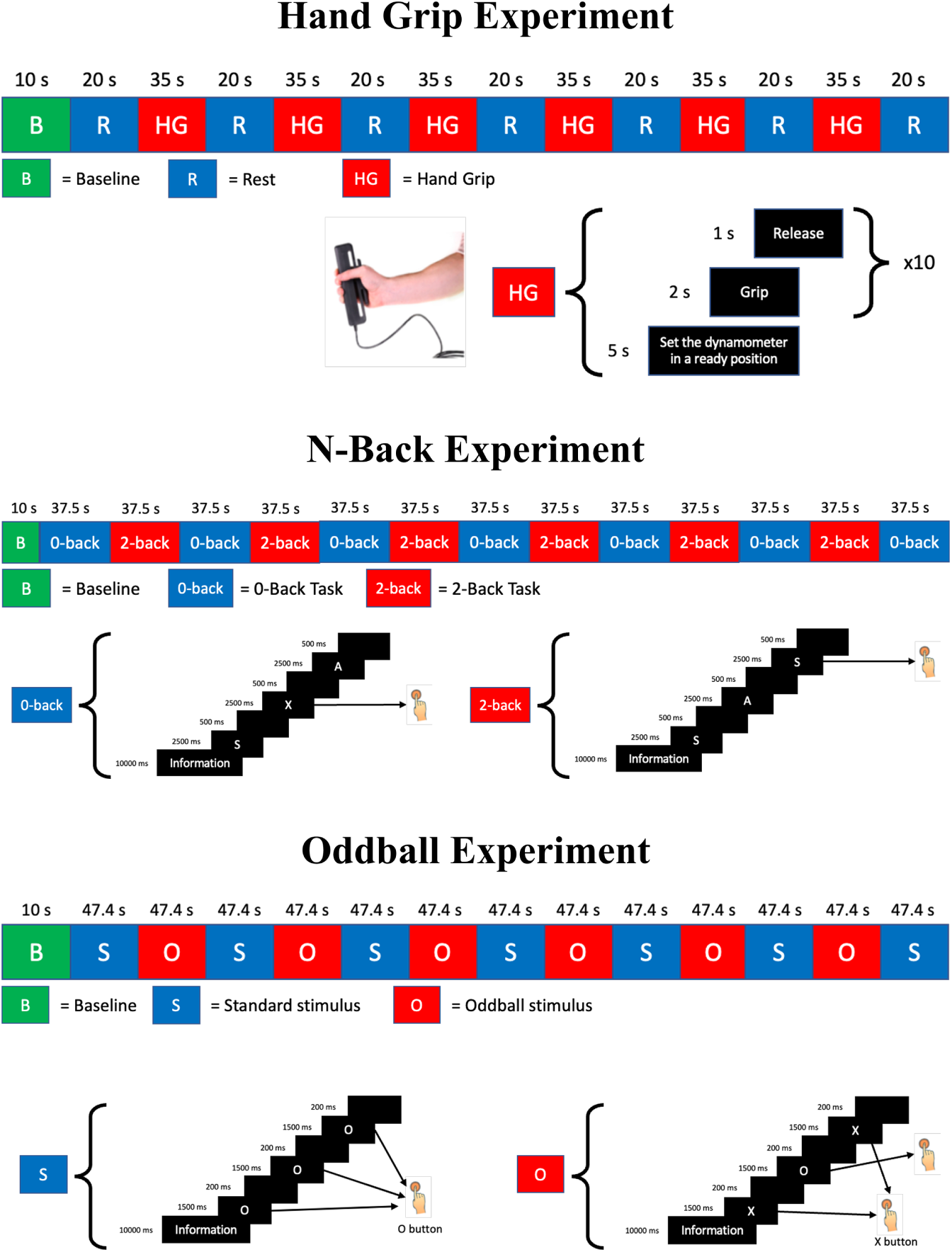
Block designs of the hand grip, N-Back, and Oddball experiments. Each experiment began with a baseline period followed by alternating task and control blocks. The hand grip task included rest and hand grip periods; the N-Back task included 0-back and 2-back conditions; and the Oddball task included standard and oddball stimulus blocks. B: baseline; R: rest; HG: hand grip; S: standard stimulus; O: oddball stimulus.

### 2.3. Data Acquisition

During the grip strength experiment, fNIRS, EMG, and hand dynamometer data were collected from participants. For the N-Back and Oddball tasks, only fNIRS data was recorded. The fNIRS data was collected using the NIR-Sport2 (NIRx Medical Technologies, LLC) device at wavelengths of 760 and 850 nm with a 10 Hz sampling rate. The device’s optodes were positioned on the scalp according to the EEG 10-10 system, with corresponding brain regions shown in Figure 2 (left). The anatomical locations of the optodes were determined using the fNIRS Optodes’ Location Decider (fOLD) application and the LONI atlas, set at a 50% specificity level (Zimeo Morais et al., 2018). EMG and hand dynamometer data from participants’ right arms were recorded using Biopac MP36 device and Biopac Student Laboratory software, with EMG electrode placement shown in Figure 2 (right).

**Figure 2:**
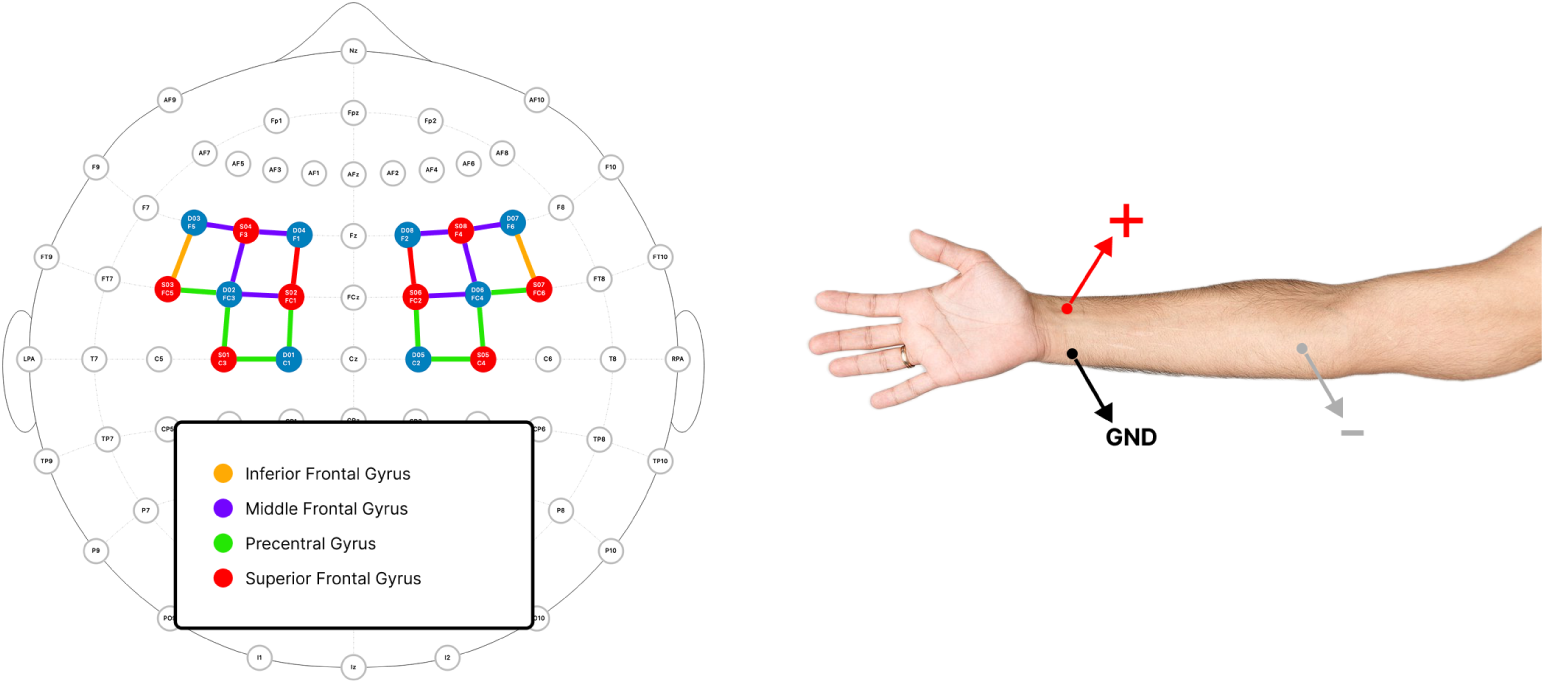
Optode and electrode placement for data acquisition. Left: fNIRS optode configuration over the frontal and motor cortices, mapped according to the 10–10 system. Channels were positioned to cover the inferior frontal gyrus, middle frontal gyrus, superior frontal gyrus, and precentral gyrus. Right: Surface EMG electrode placement on the forearm, with active (positive), reference (negative), and ground (GND) electrodes indicated.

### 2.4. Preprocessing

#### 2.4.1. fNIRS preprocessing

Raw intensity data obtained from the fNIRS device and stimulus triggers were saved in Shared Near Infrared File (SNIRF) format and preprocessed using MNE-NIRS, a Python library (Luke et al., 2021). The raw near-infrared light intensity data was first converted to optical density (OD) data. Following this, the quality of the recorded data was assessed by calculating the Scalp Coupling Index (SCI) through inter-channel correlation (El-Baba and Schury, 2025). Channels with an SCI value below 0.5 were excluded from the study, as they were deemed to have insufficient signal quality. OD data of the remaining channels are converted to oxyhemoglobin (ΔHbO) and deoxyhemoglobin (ΔHbR) concentration change data using Modified Beer Lambert Law (Cope et al., 1988). To eliminate heart rate (>1 Hz), respiratory (0.15-0.4 Hz) and Mayer wave (∼0.1 Hz) artifacts from the concentration data (Fekete et al., 2011), a 50th-order Butterworth filter with a frequency range of 0.005–0.1 Hz was applied (Yücel et al., 2016). Additionally, the Correlation-Based Signal Improvement (CBSI) was used to correct motion artifacts in the signal (Cui et al., 2010). For statistical analysis, General Linear Model (GLM) was applied to the fNIRS data (Von Lühmann et al., 2020). GLM is represented as in Equation 1:

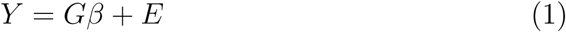

In this equation, Y represents the recorded fNIRS signal. G is the design matrix that represents the expected hemodynamic response. E represents the noise and other artifacts in the signal. *β* is a coefficient that represents the compatibility between the given stimulus and the fNIRS signal. To obtain the *β* coefficients, the equation is solved as Equation 2:

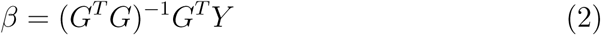

The obtained *β* coefficients were used to represent the fNIRS signal in further statistical analysis.

fNIRS signals were divided into epochs according to the experimental stimuli. During this process, baseline correction was performed by subtracting the average signal from 2 seconds of pre-stimulus period. To obtain the signals representing the brain regions shown in Figure 3 (left), the average of the signals from the channels corresponding to those regions was taken.

#### 2.4.2. EMG and hand dynamometer preprocessing

The EMG data recorded during the grip experiment at a sampling frequency of 1000 Hz which was down sampled to a frequency of 250 Hz using anti-aliasing down-pass filtering (Ortega et al., 2020). Then, a 17th-order Butterworth high-pass filter with a cutoff frequency of 110 Hz was applied to remove noise from the signal. The rectification of the signal was performed by taking the absolute value of the filtered signal. To make the muscle activities in the EMG signal, which exhibits high-frequency oscillations, more prominent, the Hilbert envelope was applied.

The hand dynamometer data was also down sampled from 1000 Hz to 250 Hz using anti-aliasing down-pass filtering. Subsequently, the Hilbert envelope was applied to smooth the signal. Both signals were segmented into epochs corresponding to the grip experiment.

### 2.5. Statistical analysis

For statistical analysis, independent samples t-test and Pearson’s correlation analysis were employed. These analyses were performed on the *β* coefficients derived from fNIRS data, the mean and maximum values computed from EMG data, and the clinical data collected prior to the experiments, such as age, hand grip strength, and vitamin D levels. The t-tests and chi-squared tests were used to analyze these variables between sarcopenic and control groups, while correlation was employed to examine potential relationships among the variables.

In addition, a 2x2 Repeated Measures ANOVA was conducted on the *β* coefficients obtained from the fNIRS data to analyze differences within conditions and between groups for each experimental task. This analysis allowed for the examination of main effects and interaction effects between factors such as group (Sarcopenic vs. Control) and experimental conditions. For the grip experiment, the ANOVA utilized a 2 (Group: Sarcopenic x Control) x 2 (Condition: Rest x Grip) design to explore both group-based differences and changes in brain activity between the resting and gripping conditions. For the N-Back experiment, a 2 (Group: Sarcopenic x Control) x 2 (Condition: 0-Back x 2-Back) design was employed to assess differences in cognitive task performance and working memory load and for the Oddball experiment, a 2 (Group: Sarcopenic x Control) x 2 (Condition: Standard Stimulus x Oddball Stimulus) design was used to investigate neural responses to attention-related stimuli.

Finally, the correlation between clinical data (grip strength and CST) and *β* coefficients were calculated to investigate the relationship between physical ability and neural activity. Additionally, correlations between the three experiments were calculated to assess the interrelations between neural responses to different tasks. In doing so, we aimed to determine whether a shared neural substrate underlies motor execution, working memory and attentional processing, thereby providing insight into the integrated neurocognitive framework that may be affected by sarcopenia.

## 3. Results

### 3.1. Clinical data results

The independent samples t-test was performed to evaluate differences between control and sarcopenic groups for numerical clinical data (Table 1). Only significant difference was observed in grip strength between sarcopenic and healthy control group (t(66) = 4.065, p < 0.001). Chi-square (*x*^2^) test was conducted to assess differences in categorical clinical data (Table 2). No significant differences were observed in the categorical clinical data.

**Table 2:** Chi-square test results for categorical clinical data.

| Variable | $\chi^2$ value | df | p-value |
| --- | --- | --- | --- |
| Gender | 0.080 | 1 | 0.778 |
| Smoking | 0.027 | 1 | 0.869 |
| Alcohol Consumption | 1.286 | 1 | 0.257 |

#### 3.1.1. EMG and hand dynamometer results

The group averages for EMG and dynamometer data are presented in Figure 3. The results of the t-test conducted on the average and maximum EMG and dynamometer values are provided in Table 3. Significant differences between both groups were found for mean EMG, mean dynamometer force and peak dynamometer force. Box plots for EMG and dynamometer for both groups are shown in Figure 4.

#### 3.1.2. ANOVA Results

Comprehensive ANOVA results for the Hand Grip, N-Back, and Oddball tasks are reported in Online Resource 1 (Tables S1–S3).

For hand grip experiment, there were no significant group main effect and interaction between group and condition was observed. A significant main effect of Condition was observed in left PcG [S1-D1: F(1,55) = 16.306, p < 0.001, S1-D2: F(1,81) = 11.577, p = 0.001, S2-D1: F(1,65) = 8.161, p = 0.006, S3-D2: F(1,72) = 6.013, p = 0.017], left MFG [S2-D2: F(1,72) = 13.729, p < 0.001, S4-D2: F(1,82) = 11.993, p = 0.001, S4-D3: F(1,77) = 17.411, p < 0.001, S4-D4: F(1,96) = 9.258, p = 0.003]. left SFG [S2-D4: F(1,81) = 5.155, p = 0.026], right PcG [S5-D5: F(1,66) = 60.722, p < 0.001, S5-D6: F(1,84) = 33.192, p < 0.001, S6-D5: F(1,70) = 53.608, p < 0.001, S7-D6: F(1,69) = 4.399, p = 0.040], right MFG [S6-D6: F(1,69) = 21.731, p < 0.001, S8-D6: F(1,82) = 30.662, p < 0.001, S8-D7: F(1,73) = 19.990, p < 0.001, S8-D8: F(1,86) = 15.960, p < 0.001], right SFG [S6-D8: F(1,70) = 14.294, p < 0.001], right IFG [S7-D7: F(1,69) = 16.300, p < 0.001] with higher activation during grip condition compared to rest.

**Table 3:** EMG and hand dynamometer t-test results between groups.

| Feature | Mean±SD Values | | Statistical Test | |
| --- | --- | --- | --- | --- |
|  | Control | Sarcopenic | t-test | p-value |
| EMG Mean | 0.81±0.45 | 0.62±0.25 | 2.135 | 0.037* |
| EMG Max. | 1.17±0.60 | 0.93±0.34 | 1.976 | 0.053 |
| Dynamometer Mean | 69.54±30.25 | 50.44±12.31 | 3.361 | 0.001*** |
| Dynamometer Max. | 96.56±35.42 | 75.51±19.66 | 2.853 | 0.006** |

Notes: \* $p < 0.05$ , \*\* $p < 0.01$ , \*\*\* $p < 0.001$ .

**Figure 3:**
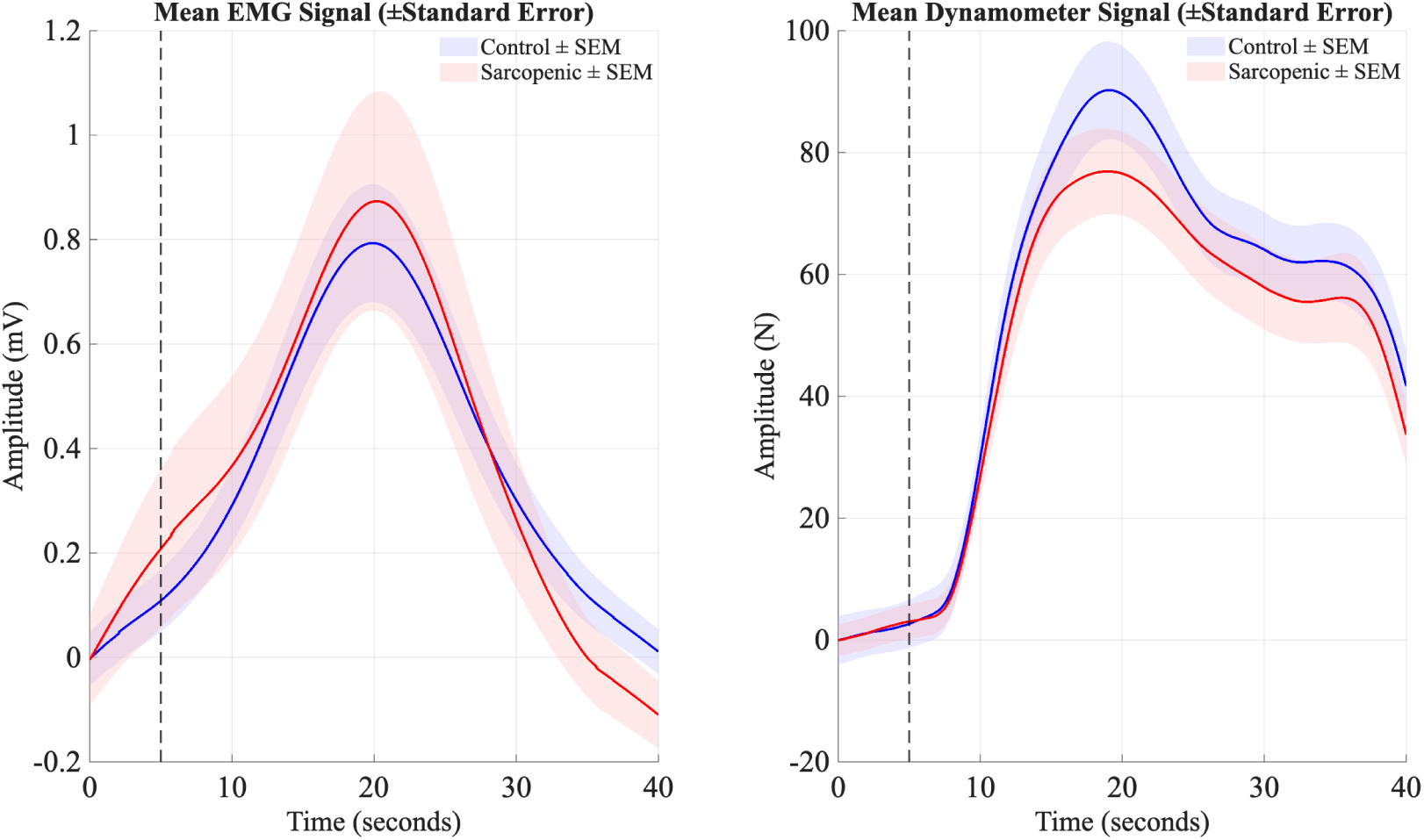
Group-averaged EMG and hand dynamometer signals during the hand grip task. Left: Mean EMG amplitude (mV) over time for control (blue) and sarcopenic (red) groups. Right: Mean dynamometer force (N) over time for both groups. Shaded areas represent the standard error. The dashed vertical line indicates the onset of the hand grip period.

**Figure 4:**
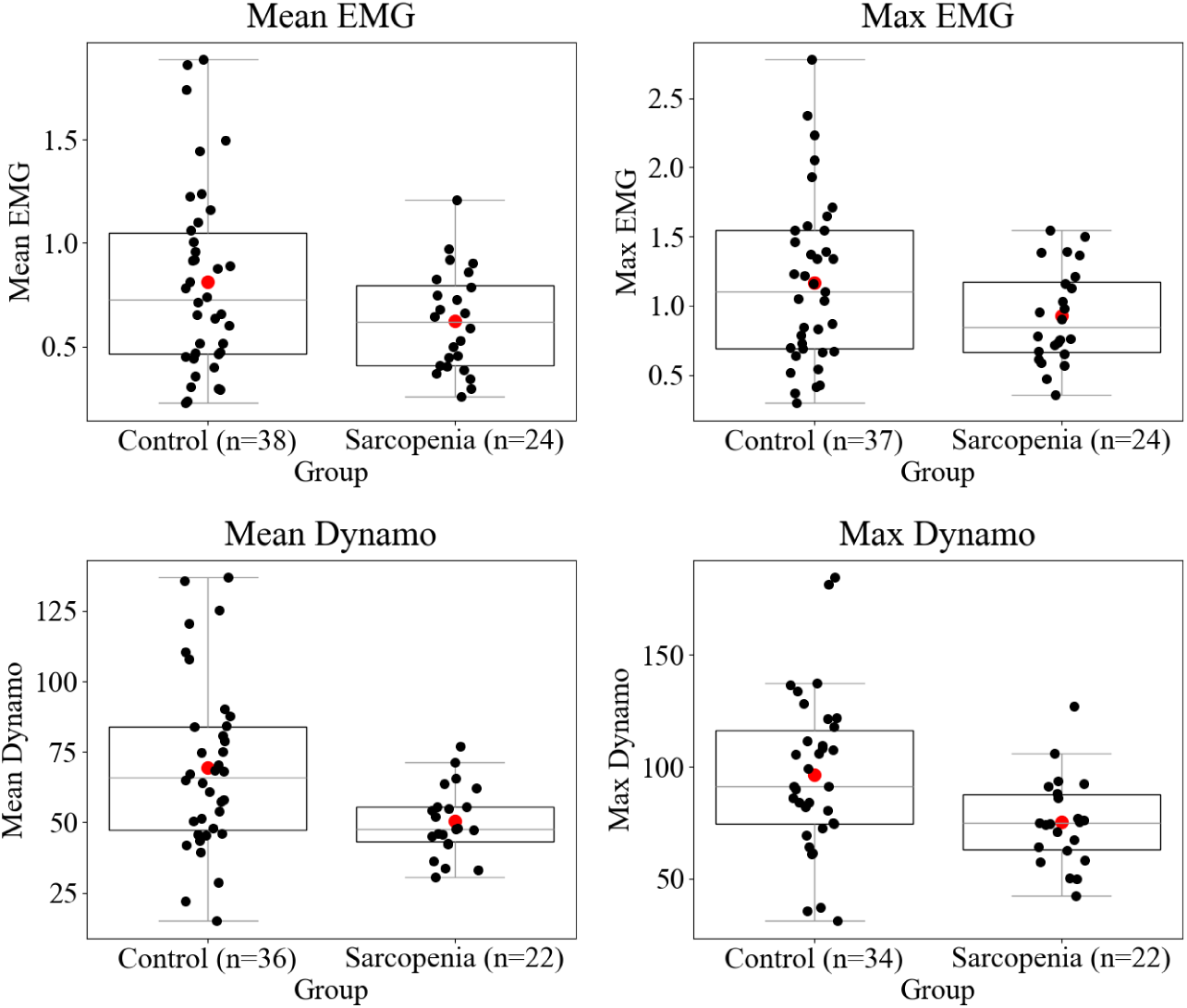
Group comparisons of EMG and hand dynamometer metrics. Top row: Mean and maximum EMG amplitude. Bottom row: Mean and maximum dynamometer force. Box plots represent the interquartile range (IQR) with the median shown as a horizontal line; whiskers indicate the range. Black dots represent individual participants and red dots indicate group means.

For the N-Back experiment, there were no significant condition effect or group x condition interaction. Significant group effects were observed in left PcG [S1-D1: F(1, 92) = 5.627, p = 0.020], left SFG [S2-D4: F(1, 101) = 6.922, p = 0.010], left MFG [S4-D3: F(1, 99) = 3.352, p = 0.070], right PcG [S5-D5: F(1,91) = 6.410, p = 0.013, S6-D5: F(1,89) = 5.041, p = 0.027], right MFG [S6-D6: F(1,95) = 6.944, p = 0.010] with higher activation in healthy control group compared to sarcopenic group. Group means for fNIRS channels with significant group effects are presented in Figure 5

In the Oddball experiment, left PcG [S1-D1: F(1,113) = 5.528, p = 0.020], right PcG [S5-D5: F(1,109) = 19.104, p < 0.001] showed significant condition differences with higher activation in oddball stimulus compared to standard stimulus. Right PcG [S6-D5: F(1,107) = 5.139, p = 0.025] and right MFG [S8-D7: F(1,113) = 6.081, p = 0.015] displayed significant group results with higher activation in sarcopenic group compared to healthy control. Right PcG [S7-D6: F(1,112) = 8.900, p = 0.004] had significant group x condition interactions. Group means for fNIRS channels with significant group effects are presented in Figure 6.

#### 3.1.3. Correlation results

When examining the correlation between the *β* coefficients obtained from the GLM analysis and clinical data, significant correlations were observed between CST scores and various brain regions. No significant correlation were found between EMG and fNIRS activity

For the grip strength task, a significant positive correlation was found in the left superior frontal gyrus (SFG) (S6-D8: r=0.279, p=0.016). In the N-back task, significant positive correlations were identified in the left precentral gyrus (PcG) (S1-D2: r=0.300, p=0.009) and the left inferior frontal gyrus (IFG) (S3-D3: r=0.297, p=0.008). Similarly, in the Oddball task, a positive correlation was observed in the left precentral gyrus (PcG) (S1-D1: r=0.232, p=0.047, S2-D1: r=0.237, p=0.045).

**Figure 5:**
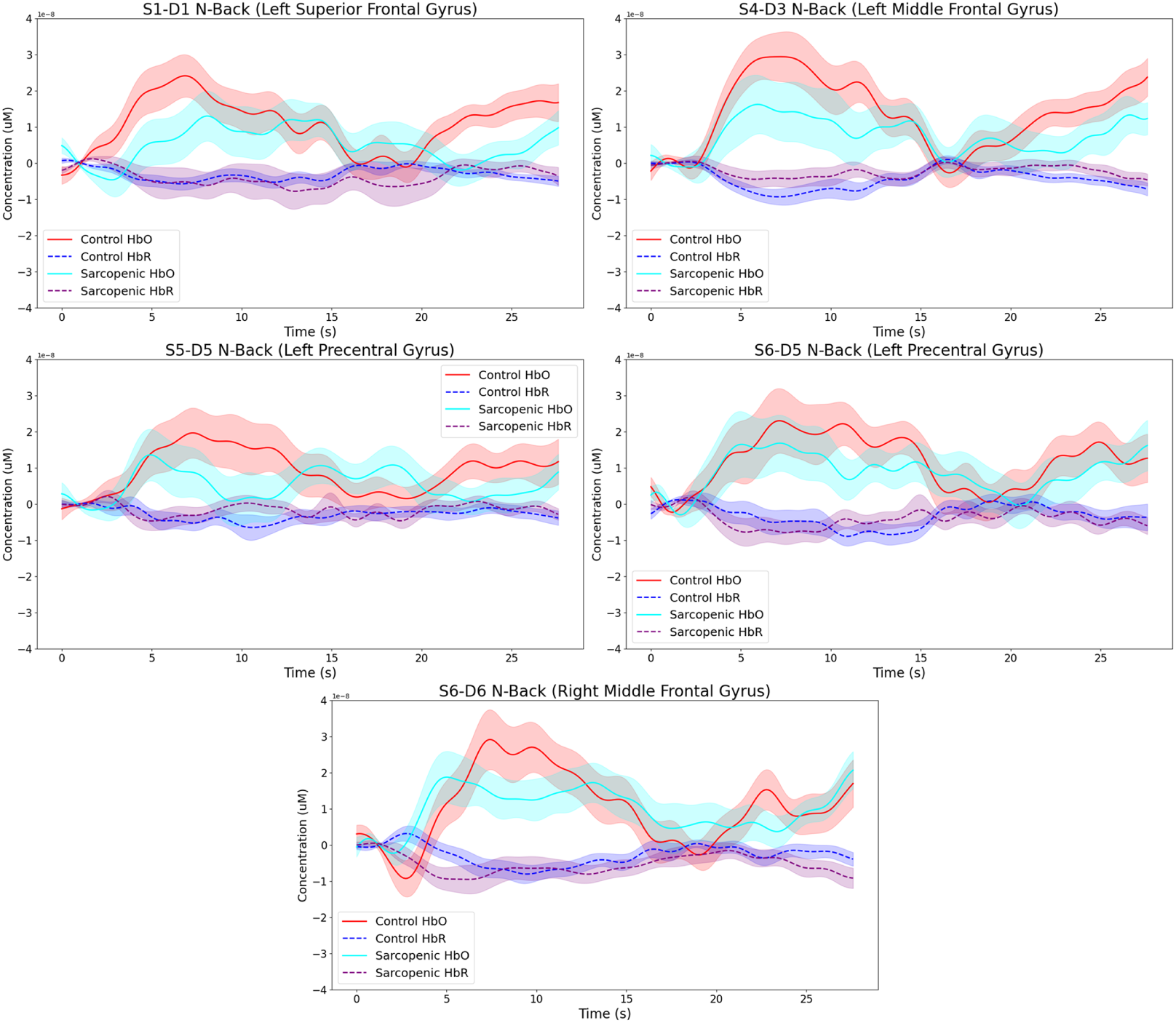
Hemodynamic responses during the N-Back task in regions showing significant group effects. Mean oxyhemoglobin (HbO; solid lines) and deoxyhemoglobin (HbR; dashed lines) concentration changes are shown for control (red/blue) and sarcopenic (cyan/purple) groups. Shaded areas represent the standard error. Panels correspond to the left superior frontal gyrus (S1-D1), left middle frontal gyrus (S4-D3), left precentral gyrus (S5-D5 and S6-D5), and right middle frontal gyrus (S6-D6).

**Figure 6:**
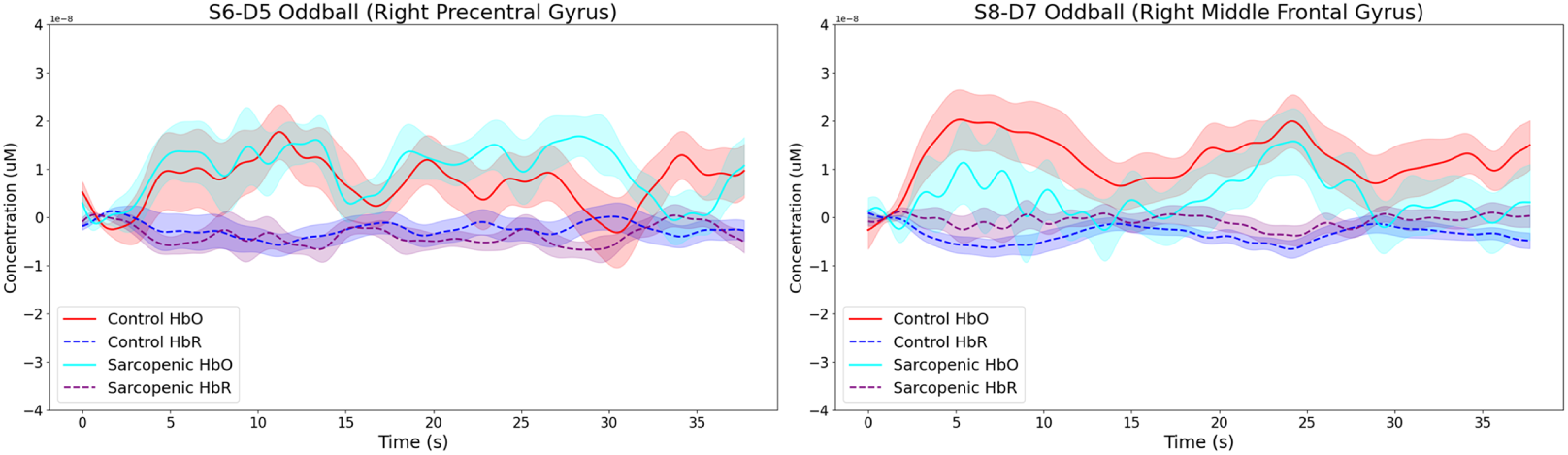
Hemodynamic responses during the Oddball task in regions showing significant group effects. Mean oxyhemoglobin (HbO; solid lines) and deoxyhemoglobin (HbR; dashed lines) concentration changes are shown for control (red/blue) and sarcopenic (cyan/purple) groups. Shaded areas represent the standard error. Panels correspond to the right pre-central gyrus (S6-D5) and right middle frontal gyrus (S8-D7).

Significant positive correlations were found between pre-experiment grip strength and specific brain regions during different tasks. In the hand grip experiment, a positive correlation was observed in the right middle frontal gyrus (MFG) (S8-D7: r=0.229, p=0.048). For the N-back task, a significant correlation was found in the right precentral gyrus (PcG) (S5-D6: r=0.320, p=0.005), and in the Oddball task, a correlation was observed in the right PcG (S7-D6: r=0.306, p=0.006).

When examining the correlations between different tasks, significant correlations were found between the hand grip experiment and the N-back experiment in the left precentral gyrus (PcG) (S1-D1: r=-0.257, p=0.027), the left middle frontal gyrus (MFG) (S4-D2: r=0.344, p=0.003, S4-D3: r=0.240, p=0.046), the right MFG (S6-D6: r=0.236, p=0.046, S8-D6: r=0.285, p=0.013), and the right superior frontal gyrus (SFG) (S6-D8: r=0.352, p=0.002).

Additionally, significant correlations were observed between the hand grip and Oddball experiments in the left PcG (S1-D1, S3-D2) and the left SFG (S2-D4) regions.

## 4. Discussion

In this study, we investigated the potential neurological markers of sarcopenia disease. We have demonstrated the effect of sarcopenia on brain function using fNIRS. Statistical analysis results revealed cognitive tasks (N-back and Oddball) significant differences between sarcopenia and healthy control groups however hand-grip task showed no significant difference between groups. These findings are consistent with recent studies emphasizing neurological underpinnings associated with sarcopenia.

T-test results for EMG and hand dynamometer data showed significant differences between groups with larger muscle activity and force output in control group compared to sarcopenia group. These findings indicate that sarcopenia involves a loss of voluntary force output and reduced neuromuscular drive during contraction.

During the hand grip experiment, there were significant differences between rest and grip condition in all regions. However, there were no significant activation differences between control and sarcopenia groups. In contrast, EMG and dynamometer t-test results demonstrated that sarcopenic individuals produced significantly lower force output and muscle activity. This dissociation suggests that, in sarcopenia, the motor cortex is engaged comparably to healthy adults yet deficits in muscle quality or neuromuscular transmission limit actual force production. Such findings support the view that strength loss in sarcopenia reflects peripheral impairments more than a failure of cortical recruitment. This interpretation is supported by Trost et al. (2023), who found that during simple motor tasks, older adults with low muscle strength exhibited similar brain activation to non-dynapenic peers (Trost et al., 2023).

During the N-back experiment, healthy control group exhibited significantly higher activation in left and right PcG, left and right MFG and left SFG regions compared to sarcopenic group. This differential activation may indicate that sarcopenia is associated with impaired cognitive function, particularly in domains related to working memory and attention. The reduced activation in the PcG within the sarcopenic group could be attributed to diminished motor control and neuromuscular efficiency, potentially affecting response execution during the button-pressing task. Previous research has shown that sarcopenic individuals exhibit increased motor reaction time and slower motor execution due to neuromuscular deficits (Pereira Da Silva Alves et al., 2023). Given the PcG’s role in initiating voluntary movement (Banker and Tadi, 2025), these findings suggest that reduced cortical activity in PcG region may contribute to the observed delays in response execution. Differences in MFG and SFG regions might indicate impairments in working memory and attention in sarcopenic subjects. Previous research has shown that sarcopenia is associated with cognitive impairments, particularly in executive function and attention-related tasks (Xing et al., 2023). Given the role of the MFG and SFG in working memory and attention (El-Baba and Schury, 2025; Boisgueheneuc et al., 2006), the observed deficits could be linked to impaired frontal lobe activity in sarcopenic individuals, leading to diminished working memory and attention control. Notably, however, there was no significant difference between 0-Back and 2-Back conditions. This lack of load-dependent activation may suggest that increasing working memory demand in 2-Back may not have been sufficient to elicit measurable changes in fNIRS activity.

In contrast to the N-back experiment, the sarcopenic group exhibited significantly higher activation in right PcG and right MFG during the Oddball experiment compared to healthy control group. This increased activation may reflect a compensatory neural response due to deficits in attention and response inhibition. Similar compensatory functional changes in aging populations have also been reported by Goh and Park, who demonstrated that despite structural decline, older adults maintain cognitive performance by recruiting additional neural resources (Goh and Park, 2009).The Oddball task requires attentional shifting and response execution, processes that rely on top-down executive control from the MFG and motor preparation in the PcG. Given the sarcopenic group’s known impairments in reaction time and executive function, they may have recruited additional neural resources in the right hemisphere to compensate for their cognitive and motor limitations (Clément et al., 2013). This lateralized activation pattern aligns with prior research suggesting that the right hemisphere plays a dominant role in attentional control and response monitoring (Fassbender et al., 2004). The lack of significant differences in left hemisphere regions further supports the idea that the right MFG and PcG were specifically engaged to compensate for attentional demands. In contrast, the control group may have executed the task more efficiently, requiring less neural activation in these regions. These results suggest that in low cognitive load tasks like Oddball, sarcopenic individuals may still be able to engage compensatory mechanisms, whereas in higher cognitive load tasks like N-back, these mechanisms may be insufficient, leading to reduced bilateral activation (Staffen et al., 2002).

The correlation analyses revealed significant relationships between brain activity and clinical measures, highlighting the interplay between motor and cognitive functions. The positive correlation between HGS and right MFG activity during the hand grip experiment suggests that individuals with greater grip strength may recruit higher-order cognitive processes to regulate and sustain force output (Davis et al., 2022). Given the role of the MFG in executive functions, this finding implies that stronger individuals may engage cognitive control mechanisms to optimize motor performance (El-Baba and Schury, 2025; Carson, 2018). Moreover, the significant positive correlations between HGS and right PcG activity in N-back and Oddball experiments indicate that individuals with greater grip strength exhibit higher motor-related activation even during cognitive tasks. This suggests that motor function and cognitive processing share common neural pathways (Sato et al., 2024).

Similarly, CST scores correlated positively with left PcG activation in the N-back and Oddball experiments, suggesting that individuals with better mobility exhibit stronger motor-cognitive interaction even in cognitively demanding tasks (Rosano et al., 2005). This supports the idea that lower-limb strength and neuromuscular function are linked to cognitive-motor efficiency (Gnosa et al., 2019). Additionally, the positive correlation between CST scores and left IFG activation in the N-back experiment suggests that better mobility may be associated with enhanced inhibitory control during cognitive tasks (Georgantas et al., 2024). The significant correlation between CST scores and left SFG activation during the hand grip experiment suggests that individuals with better mobility rely more on high-order cognitive control mechanisms during motor execution. Since SFG is involved in executive function and attentional regulation, this may indicate that higher-functioning individuals engage cognitive resources more efficiently to support motor performance (Gnosa et al., 2019).

The correlation analysis also revealed significant relationships between brain activity across the hand grip, N-back, and Oddball experiments, suggesting shared neural substrates underlying motor execution, working memory, and attentional control (Ikkai and Curtis, 2011). Notably, left PcG activity showed a negative correlation, whereas left and right MFG and right SFG activity exhibited positive correlations between the hand grip and N-back tasks. This pattern suggests that while basic motor execution (PcG) is task-dependent and may be recruited differently based on the cognitive demands of the task, executive control regions (MFG, SFG) are consistently engaged across both motor and cognitive functions (Gerver et al., 2020). Furthermore, left PcG and SFG activity correlated positively between the hand grip and Oddball tasks, reinforcing the idea that motor and attentional processes share overlapping neural networks (Larson-Prior et al., 2009). Unlike the N-back task, which primarily relies on working memory without immediate motor demands, the Oddball task requires rapid responses to infrequent stimuli, engaging both motor and cognitive control mechanisms simultaneously (Zhao et al., 2023). This distinction may explain why motor-related activation (PcG) correlated positively between grip and Oddball tasks, reflecting a stronger interaction between motor and attentional control processes.

## 5. Conclusion

This study highlights the neurocognitive impact of sarcopenia, demonstrating its association with altered brain activity in motor and cognitive control regions. The findings suggest that sarcopenia is a musculoskeletal condition and also involves significant changes in neural activation patterns, particularly in areas related to motor control, working memory and attentional processing. These results emphasize the need to consider both cognitive and neuromuscular factors when assessing sarcopenia-related functional decline.

The observed compensatory neural mechanisms in sarcopenic individuals suggest potential targets for interventions aimed at preserving motor and cognitive function. Future research should explore longitudinal studies to track neurocognitive changes over time and investigate the efficacy of combined motor and cognitive training programs in mitigating sarcopenia-related impairments.

By integrating neuroimaging techniques such as fNIRS with traditional clinical assessments, this study contributes to a more comprehensive understanding of sarcopenia, paving the way for early detection and more effective therapeutic strategies.

## Ethics approval

The study was approved by the TOBB University of Economics and Technology Human Research Ethics Committee (Protocol No. 2022-25). Written informed consent was obtained from all participants prior to data collection.

## Data availability

The datasets generated and analyzed during the current study are not publicly available due to participant confidentiality but are available from the corresponding author on reasonable request.

## Code availability

The analysis code supporting the findings of this study is available from the corresponding author upon reasonable request.

## Author contributions (CRediT)

B.M.Ş.: Conceptualization, Methodology, Data curation, Formal analysis, Visualization, Writing - original draft. A.E.: Supervision, Methodology, Writing - review & editing. K.E., M.E.D., Ö.K.: Investigation, Resources. B.K., M.K.: Validation, Writing - review & editing. All authors approved the final manuscript.

## Acknowledgements

The authors thank all participants for their voluntary involvement in this study. We also acknowledge the support and clinical collaboration provided by the team at Dr. Abdurrahman Yurtaslan Ankara Oncology Training and Research Hospital during data collection. This work was supported by the Scientific and Technological Research Council of Türkiye (TÜBİTAK) under Project No. 122E210 within the TÜBİTAK 1001 program.

