## Supplementary Tables 1-3 for "Neural Indicators of Motor and Cognitive Functioning in Sarcopenia Using Functional Near-Infrared Spectroscopy"

### Online Resource 1. Supplementary Tables

**Table S1:** Grip experiment ANOVA results.

| Channel | Region | Condition | Group | Group x Condition |
|---|---|---|---|---|
| S1 - D1 | PcG (Left) | F(1,55) = 16.306, p < 0.001* | F(1,55) = 0.002, p = 0.958 | F(1,55) = 7.079, p = 0.010* |
| S1 - D2 | PcG (Left) | F(1,81) = 11.577, p = 0.001* | F(1,81) = 0.019, p = 0.890 | F(1,81) = 0.235, p = 0.629 |
| S2 - D1 | PcG (Left) | F(1,65) = 8.161, p = 0.006* | F(1,65) = 1.202, p = 0.277 | F(1,65) = 10.055, p = 0.002* |
| S2 - D2 | MFG (Left) | F(1,72) = 13.729, p < 0.001* | F(1,72) = 0.239, p = 0.626 | F(1,72) = 0.006, p = 0.938 |
| S2 - D4 | SFG (Left) | F(1,81) = 5.155, p = 0.026* | F(1,81) = 0.510, p = 0.477 | F(1,81) = 1.702, p = 0.196 |
| S3 - D2 | PcG (Left) | F(1,72) = 6.013, p = 0.017* | F(1,72) = 0.269, p = 0.605 | F(1,72) = 2.925, p = 0.092 |
| S3 - D3 | IFG (Left) | F(1,75) = 3.326, p = 0.072 | F(1,75) = 0.028, p = 0.867 | F(1,75) = 0.384, p = 0.537 |
| S4 - D2 | MFG (Left) | F(1,82) = 11.993, p = 0.001* | F(1,82) = 0.691, p = 0.408 | F(1,82) = 0.453, p = 0.503 |
| S4 - D3 | MFG (Left) | F(1,77) = 17.411, p < 0.001* | F(1,77) = 0.333, p = 0.565 | F(1,77) = 0.233, p = 0.631 |
| S4 - D4 | MFG (Left) | F(1,96) = 9.258, p = 0.003* | F(1,96) = 0.001, p = 0.972 | F(1,96) = 0.523, p = 0.471 |
| S5 - D5 | PcG (Right) | F(1,66) = 60.722, p < 0.001* | F(1,66) = 0.767, p = 0.384 | F(1,66) = 0.017, p = 0.895 |
| S5 - D6 | PcG (Right) | F(1,84) = 33.192, p < 0.001* | F(1,84) = 0.385, p = 0.536 | F(1,84) = 1.346, p = 0.249 |
| S6 - D5 | PcG (Right) | F(1,70) = 53.608, p < 0.001* | F(1,70) = 1.361, p = 0.247 | F(1,70) = 0.000, p = 0.991 |
| S6 - D6 | MFG (Right) | F(1,69) = 21.731, p < 0.001* | F(1,69) = 0.181, p = 0.671 | F(1,69) = 0.013, p = 0.908 |
| S6 - D8 | SFG (Right) | F(1,70) = 14.294, p < 0.001* | F(1,70) = 0.007, p = 0.931 | F(1,70) = 0.771, p = 0.383 |
| S7 - D6 | PcG (Right) | F(1,69) = 4.399, p = 0.040* | F(1,69) = 0.266, p = 0.607 | F(1,69) = 0.502, p = 0.481 |
| S7 - D7 | IFG (Right) | F(1,69) = 16.300, p < 0.001* | F(1,69) = 2.062, p = 0.155 | F(1,69) = 0.107, p = 0.743 |
| S8 - D6 | MFG (Right) | F(1,82) = 30.662, p < 0.001* | F(1,82) = 0.045, p = 0.832 | F(1,82) = 3.297, p = 0.073 |
| S8 - D7 | MFG (Right) | F(1,73) = 19.990, p < 0.001* | F(1,73) = 0.000, p = 0.991 | F(1,73) = 0.674, p = 0.414 |
| S8 - D8 | MFG (Right) | F(1,86) = 15.960, p < 0.001* | F(1,86) = 1.936, p = 0.168 | F(1,86) = 0.342, p = 0.560 |

** p < 0.05*

**Table S2:** N-Back experiment ANOVA results.

| Channel | Region | Condition | Group | Group x Condition |
|---|---|---|---|---|
| S1 - D1 | PcG (Left) | F(1,92) = 0.216, p = 0.643 | F(1,92) = 5.627, p = 0.020* | F(1,92) = 0.260, p = 0.611 |
| S1 - D2 | PcG (Left) | F(1,109) = 0.055, p = 0.815 | F(1,109) = 1.617, p = 0.206 | F(1,109) = 3.628, p = 0.059 |
| S2 - D1 | PcG (Left) | F(1,89) = 0.946, p = 0.333 | F(1,89) = 0.124, p = 0.725 | F(1,89) = 1.892, p = 0.172 |
| S2 - D2 | MFG (Left) | F(1,98) = 1.309, p = 0.255 | F(1,98) = 0.737, p = 0.393 | F(1,98) = 0.025, p = 0.873 |
| S2 - D4 | SFG (Left) | F(1,101) = 0.266, p = 0.607 | F(1,101) = 6.922, p = 0.010* | F(1,101) = 0.035, p = 0.851 |
| S3 - D2 | PcG (Left) | F(1,99) = 0.049, p = 0.824 | F(1,99) = 3.383, p = 0.069 | F(1,99) = 0.337, p = 0.563 |
| S3 - D3 | IFG (Left) | F(1,105) = 2.852, p = 0.094 | F(1,105) = 1.498, p = 0.224 | F(1,105) = 2.976, p = 0.087 |
| S4 - D2 | MFG (Left) | F(1,105) = 0.273, p = 0.602 | F(1,105) = 2.640, p = 0.107 | F(1,105) = 0.115, p = 0.735 |
| S4 - D3 | MFG (Left) | F(1,99) = 3.352, p = 0.070 | F(1,99) = 5.064, p = 0.027* | F(1,99) = 3.125, p = 0.080 |
| S4 - D4 | MFG (Left) | F(1,109) = 0.224, p = 0.637 | F(1,109) = 2.097, p = 0.150 | F(1,109) = 1.514, p = 0.221 |
| S5 - D5 | PcG (Right) | F(1,91) = 1.617, p = 0.207 | F(1,91) = 6.410, p = 0.013* | F(1,91) = 1.352, p = 0.248 |
| S5 - D6 | PcG (Right) | F(1,97) = 2.108, p = 0.150 | F(1,97) = 0.252, p = 0.616 | F(1,97) = 0.655, p = 0.420 |
| S6 - D5 | PcG (Right) | F(1,89) = 2.671, p = 0.106 | F(1,89) = 5.041, p = 0.027* | F(1,89) = 0.674, p = 0.414 |
| S6 - D6 | MFG (Right) | F(1,95) = 1.096, p = 0.298 | F(1,95) = 6.944, p = 0.010* | F(1,95) = 0.037, p = 0.848 |
| S6 - D8 | SFG (Right) | F(1,96) = 0.007, p = 0.930 | F(1,96) = 0.105, p = 0.746 | F(1,96) = 0.452, p = 0.503 |
| S7 - D6 | PcG (Right) | F(1,95) = 0.706, p = 0.403 | F(1,95) = 0.660, p = 0.418 | F(1,95) = 1.900, p = 0.171 |
| S7 - D7 | IFG (Right) | F(1,85) = 2.916, p = 0.091 | F(1,85) = 0.372, p = 0.543 | F(1,85) = 0.551, p = 0.460 |
| S8 - D6 | MFG (Right) | F(1,106) = 0.337, p = 0.563 | F(1,106) = 3.005, p = 0.086 | F(1,106) = 0.015, p = 0.902 |
| S8 - D7 | MFG (Right) | F(1,101) = 0.131, p = 0.718 | F(1,101) = 2.581, p = 0.111 | F(1,101) = 0.022, p = 0.880 |
| S8 - D8 | MFG (Right) | F(1,97) = 2.949, p = 0.089 | F(1,97) = 0.528, p = 0.469 | F(1,97) = 1.375, p = 0.244 |

* $p < 0.05$

**Table S3.** Oddball experiment ANOVA results.

| Channel | Region | Condition | Group | Group x Condition |
|---|---|---|---|---|
| S1 - D1 | PcG (Left) | F(1,113) = 5.528, p = 0.020* | F(1,113) = 2.026, p = 0.157 | F(1,113) = 0.437, p = 0.510 |
| S1 - D2 | PcG (Left) | F(1,118) = 1.852, p = 0.176 | F(1,118) = 2.450, p = 0.120 | F(1,118) = 0.783, p = 0.378 |
| S2 - D1 | PcG (Left) | F(1,109) = 0.143, p = 0.706 | F(1,109) = 0.014, p = 0.904 | F(1,109) = 0.461, p = 0.498 |
| S2 - D2 | MFG (Left) | F(1,110) = 1.129, p = 0.290 | F(1,110) = 1.146, p = 0.287 | F(1,110) = 1.147, p = 0.287 |
| S2 - D4 | SFG (Left) | F(1,112) = 2.894, p = 0.092 | F(1,112) = 0.662, p = 0.417 | F(1,112) = 0.008, p = 0.926 |
| S3 - D2 | PcG (Left) | F(1,111) = 0.172, p = 0.679 | F(1,111) = 0.924, p = 0.338 | F(1,111) = 0.670, p = 0.415 |
| S3 - D3 | IFG (Left) | F(1,114) = 0.110, p = 0.740 | F(1,114) = 1.522, p = 0.220 | F(1,114) = 1.572, p = 0.212 |
| S4 - D2 | MFG (Left) | F(1,117) = 0.081, p = 0.776 | F(1,117) = 0.093, p = 0.761 | F(1,117) = 0.880, p = 0.350 |
| S4 - D3 | MFG (Left) | F(1,114) = 0.423, p = 0.516 | F(1,114) = 0.674, p = 0.413 | F(1,114) = 0.029, p = 0.864 |
| S4 - D4 | MFG (Left) | F(1,118) = 0.605, p = 0.438 | F(1,118) = 3.106, p = 0.081 | F(1,118) = 0.023, p = 0.878 |
| S5 - D5 | PcG (Right) | F(1,109) = 19.104, p = 0.00* | F(1,109) = 0.794, p = 0.375 | F(1,109) = 0.118, p = 0.731 |
| S5 - D6 | PcG (Right) | F(1,114) = 3.573, p = 0.061 | F(1,114) = 0.405, p = 0.525 | F(1,114) = 0.025, p = 0.873 |
| S6 - D5 | PcG (Right) | F(1,107) = 1.453, p = 0.231 | F(1,107) = 5.139, p = 0.025* | F(1,107) = 0.238, p = 0.626 |
| S6 - D6 | MFG (Right) | F(1,105) = 1.790, p = 0.184 | F(1,105) = 3.813, p = 0.054 | F(1,105) = 2.889, p = 0.092 |
| S6 - D8 | SFG (Right) | F(1,99) = 0.229, p = 0.633 | F(1,99) = 0.989, p = 0.322 | F(1,99) = 1.272, p = 0.262 |
| S7 - D6 | PcG (Right) | F(1,112) = 0.930, p = 0.337 | F(1,112) = 2.048, p = 0.155 | F(1,112) = 8.900, p = 0.004* |
| S7 - D7 | IFG (Right) | F(1,110) = 0.935, p = 0.336 | F(1,110) = 0.373, p = 0.543 | F(1,110) = 1.024, p = 0.314 |
| S8 - D6 | MFG (Right) | F(1,119) = 0.000, p = 0.990 | F(1,119) = 1.126, p = 0.291 | F(1,119) = 1.871, p = 0.174 |
| S8 - D7 | MFG (Right) | F(1,113) = 0.635, p = 0.427 | F(1,113) = 6.081, p = 0.015* | F(1,113) = 1.029, p = 0.312 |
| S8 - D8 | MFG (Right) | F(1,121) = 0.004, p = 0.949 | F(1,121) = 3.075, p = 0.082 | F(1,121) = 1.039, p = 0.310 |

* $p < 0.05$
